# Associations of Loneliness and Social Isolation with Healthspan and Lifespan in the US Health and Retirement Study

**DOI:** 10.1101/2020.07.10.20147488

**Authors:** CL Crowe, BW Domingue, G Hu, KM Keyes, Dy Kwon, DW Belsky

## Abstract

**Background:** Loneliness and social isolation are emerging public health challenges for aging populations.

**Methods:** We followed N=11,302 US Health and Retirement Study (HRS) participants aged 50-95 from 2006-2014 to measure persistence of experiences of loneliness and exposure to social isolation. We tested associations of longitudinal loneliness and social isolation phenotypes with disability, morbidity, mortality, and biological aging through 2018.

**Results:** During follow-up, 18% of older adults met criteria for loneliness, with 6% meeting criteria at two or more follow-up assessments. For social isolation, these fractions were 21% and 8%. HRS participants who experienced loneliness and were exposed to social isolation were at increased risk for disease, disability, and mortality. Those experiencing persistent loneliness were at a 57% increased hazard of mortality compared to those who never experienced loneliness. For social isolation, the increase was 28%. Effect-sizes were somewhat larger for counts of prevalent activity limitations and somewhat smaller for counts of prevalent chronic diseases. Covariate adjustment for socioeconomic and psychological risks attenuated but did not fully explain associations. Older adults who experienced loneliness and were exposed to social isolation also exhibited physiological indications of advanced biological aging (Cohen’s-d for persistent loneliness and social isolation=0.26 and 0.21, respectively). For loneliness, but not social isolation, persistence was associated with increased risk.

**Conclusion:** Deficits in social connectedness prevalent in a national sample of US older adults were associated with morbidity, disability, and mortality and with more advanced biological aging. Bolstering social connectedness to interrupt experiences of loneliness may promote healthy aging.

## INTRODUCTION

Experiences of loneliness and exposure to social isolation are prevalent among older adults, with an estimated 20-30% reporting some loneliness or social isolation (1,2). They are also associated with increased morbidity and mortality (2–7). Loneliness and social isolation therefore represent an emerging priority for public health intervention, the urgency of which is highlighted by the impact of shelter-in-place policies implemented to mitigate the COVID-19 pandemic (8–11).

Loneliness and social isolation are distinct constructs representing different aspects of social connectedness (12). Loneliness is the subjective feeling of being isolated. Social isolation is the objective state of having limited social interactions. Interventions are now being developed to reduce loneliness and social isolation with the aim of improving health and well-being among older adults (2,13–15). While there is some evidence that reducing loneliness may improve symptoms of depression in older adults (16), it is not known if intervention on loneliness and social isolation can impact physical health-related features of healthy aging.

Cross-sectional studies report associations of loneliness and social isolation with physical health deficits in older adults and also with mortality (2–4,17–19). However, cross-sectional data cannot rule out confounding of associations by pre-existing economic and psychological risk factors that may interfere with formation and maintenance of social connections and impair healthy aging. Cross-sectional studies also cannot exclude the possibility of reverse causation, in which disease and disability lead to loneliness and social isolation. A further challenge is that cross-sectional data cannot quantify the persistence of loneliness and social isolation, which may be an important dimension of their impact on healthy aging. Longitudinal data are therefore needed to address four questions about links from experiences of loneliness and exposure to social isolation to disease, disability, and mortality:

First, are risks associated with experiences of loneliness and exposure to social isolation independent of economic and psychological vulnerabilities that may cause both social disconnection and deficits in healthy aging? Household poverty and adverse neighborhood conditions can cause older people to become socially disconnected from their communities and are also associated with disease, disability, and mortality (20–22). In parallel, psychological vulnerabilities that put people at risk for loneliness and social isolation, including depressive symptoms and related personality features, are also linked with deficits in healthy aging (23–26). Measurements of economic and psychological vulnerability are needed to disentangle the effects of loneliness and social isolation on deficits in healthy aging from the effects of correlated risk factors.

Second, do experiences of loneliness and exposure to social isolation precede the onset of deficits in healthy aging or, instead, could deficits in healthy aging cause individuals to become lonely or socially isolated? Meta-analyses support deficits in social connectedness as predictors of future cardiovascular morbidity and all-cause mortality (7,27). However, data are more limited for prospective relationships with other types of morbidity and disability (2–4,28).

Longitudinal data can help clarify the extent of prospective links between deficits in social connectedness and deficits in healthy aging (17).

Third, does the persistence of experiences of loneliness and exposure to social isolation worsen health impacts? Interventions to address loneliness and social isolation aim to improve health by reducing the burden of loneliness and social isolation among individuals who are already lonely and socially isolated (13). However, it is not known if reducing the persistence of loneliness and social isolation will offer protection against deficits in healthy aging. Studies with measures of loneliness and social isolation at multiple time points can compare healthy aging outcomes among those whose symptoms persist to those with intermittent symptoms.

Fourth, there is need to identify biological measurements that can inform mechanisms through which experiences of loneliness and exposure to social isolation may impact healthy aging and provide a healthy-aging surrogate endpoint for interventions to address deficits in social connectedness. Biological aging is hypothesized to be the core mechanism driving age-dependent increases in risk for disease, disability, and mortality (29). There is already evidence that loneliness may compromise immune-system integrity, driving systemic inflammation (30), a pillar of aging (31). However, no studies have yet tested relationships of loneliness and social isolation with biological aging.

To address these questions and build knowledge to inform design of future programs and policies, we analyzed data from the US Health and Retirement Study (HRS), a large national sample of older adults followed longitudinally from 1992 and most recently surveyed in 2016-2018. HRS surveys older adults aged 50+, allowing us to investigate healthy aging sequelae of experiences of loneliness and exposure to social isolation across the second half of the life course. We conducted analysis to evaluate associations of loneliness and social isolation with three key dimensions of healthy aging, mortality, disability, and morbidity, and explored a potential link between experiences of loneliness and exposure to social isolation and more advanced biological aging.

## METHODS

### Study Population

The HRS is a longitudinal biennial cohort study of a nationally representative sample of non-institutionalized adults over the age of 50 and their spouses. The HRS selected participants using multistage probability sampling designed to represent adults over the age of 50 in the United States. We analyzed HRS data from RAND corporation (32) including 42,042 participants. We linked RAND files with data from the HRS Leave Behind Questionnaires (LBQ) collected during 2006-2014 (33) and from the HRS Venous Blood Study (VBS) collected in 2016 (34).

### Measures

#### Loneliness & Social Isolation

We measured experiences of loneliness and exposure to social isolation from data collected during 2006-2014 in the HRS Core Interview and LBQ. The LBQ is a self-administered survey about life circumstances, subjective well-being, and lifestyle. A random subsample of 50% of HRS participants completed the LBQ in 2006 and the other 50% in 2008. Thereafter, these subsamples completed the LBQ at alternating waves (i.e., every four years).

#### Loneliness

We measured experiences of loneliness using a 3-item version of the Revised UCLA (R-UCLA) Loneliness Scale (35). Participants rated how frequently they felt they were 1) lacking companionship, 2) left out, and 3) isolated from others on a 3-point scale. Previous analysis showed this version to have similar psychometric properties to the original 20-Item version (35). We coded item responses so that higher scores corresponded to more severe experiences of loneliness. To account for missing item-level data, we pro-rated scale scores for participants who responded to at least two of the three items by summing the non-missing item scores, dividing by the number of non-missing items, and multiplying by the total number of items in the scale. Final scores ranged from 3-9. We followed the procedure of Steptoe and colleagues (36) and classified participants in the top quintile of scale scores as lonely. This procedure classified participants scoring ≥7 as lonely.

#### Social Isolation

There is not yet a gold standard measure of exposure to social isolation. Consensus in the field is that scales should comprise multiple items and measure relationships with individuals, groups, and community organizations (37,38). We used a six-item scale meeting these criteria first validated in the English Longitudinal Study of Aging (ELSA) (5,36) and adapted to the HRS (39). We assigned a social isolation score to each participant based on whether they 1) were unmarried, 2) lived alone, 3) had less than monthly contact with children, 4) had less than monthly contact with other family members, 5) had less than monthly contact with friends, and 6) did not participate monthly in any groups, clubs, or other social organizations, yielding scores 0-6. We calculated scores for participants providing data for at least three of the six items. We followed the procedure used for loneliness and classified participants in the top quintile of scale scores as socially isolated. This procedure classified participants scoring ≥3 as socially isolated.

#### Persistent Exposure Classification

For loneliness and social isolation, we classified participants with scores meeting or exceeding the threshold score at two or more assessment waves as having persistent experiences of loneliness or exposure to social isolation. We classified participants meeting or exceeding the threshold at only one assessment as having intermittent experiences of loneliness or exposure to social isolation.

#### Sensitivity Analysis

We conducted sensitivity analysis to evaluate alternative measurements and thresholds to identify experiences of loneliness and exposure to social isolation (**Supplemental Methods**).

#### Deficits in Healthy Aging

Aging is the leading risk factor for many different chronic diseases and disabilities. However, not everyone experiences the onset of these conditions at the same rate. Some individuals remain free of chronic disease and maintain functioning into late life, whereas others develop chronic disease and disability much earlier (40). To understand how loneliness and social isolation relate to individual differences in healthy aging, we analyzed three dimensions of this process in parallel.

Mortality measures the lifespan dimension of healthy aging. HRS ascertained death dates for participants from linkages with the National Death Index and from reports in exit interviews and in interviews with spouses.

Disability measures the healthspan dimension of healthy aging. We measured disability from counts of Activities of Daily Living (ADL) and Instrumental Activities of Daily Living (IADL) limitations. We measured ADL disability as a count of the following activities with which the participant reported having at least some difficulty: bathing, dressing, eating, getting in/out of bed, and walking across a room. We measured IADL disability as a count of the following activities with which the participant reported having at least some difficulty: using the phone, managing money, taking medications, shopping for groceries, and preparing hot meals. We analyzed counts of ADL and IADL disability as 0-3+.

Chronic disease prevalence provides a second measure of the healthspan dimension of healthy aging. We measured chronic diseases as a count of aging-related chronic conditions participants reported having been diagnosed with by a physician. The conditions were: high blood pressure, diabetes, cancer, chronic lung disease, heart disease, and stroke. We analyzed counts of chronic disease diagnoses as 0-3+.

We measured biological aging from data collected in HRS’s 2016 Venous Blood Study (34) using the “Phenotypic Age” algorithm (41–43). There are several methods to quantify biological aging from blood chemistry data. We focused on Phenotypic Age because comparative studies suggest this measure is more predictive of mortality, disability, and morbidity as compared to leading alternatives (42,44,45). The Phenotypic Age algorithm was developed from a machine learning analysis of mortality in the National Health and Nutrition Examination Surveys (NHANES) III dataset (41). The analysis screened 42 blood chemistry biomarkers and chronological age to devise a prediction algorithm. The resulting algorithm included chronological age and nine biomarkers: albumin, alkaline phosphatase, creatinine, C-reactive protein, glucose, mean cell volume, red cell distribution width, white blood cell count, and lymphocyte percent. The algorithm produces a value denominated in the metric of years. The years correspond to the age at which an individual’s risk of death would be approximately normal in the NHANES III sample. A phenotypic age older than a person’s true chronological age indicates more advanced biological aging.

#### Social and Economic Circumstances

We measured participants’ social and economic circumstances across three domains: neighborhood conditions, household wealth, and education. We measured neighborhood conditions from 2006-2014, household wealth from 1993-2016, and education at each participant’s first HRS interview. Details are reported in the **Supplemental Methods**.

#### Psychological Vulnerabilities

We measured participants’ psychological vulnerabilities from assessments of the personality trait neuroticism and of symptoms of depression. We measured neuroticism at each participant’s first LBQ interview and symptoms of depression between their first HRS interview after 1992 and their first LBQ interview. Of the 11,302 participants in the analysis sample, we measured neuroticism in 11,172 participants and depressive symptoms in 11,251 participants. We excluded those with missing data from analysis. Details are reported in the **Supplemental Methods**.

A timeline of exposure and outcome assessments is provided in **Figure 1**.

**Figure 1.**
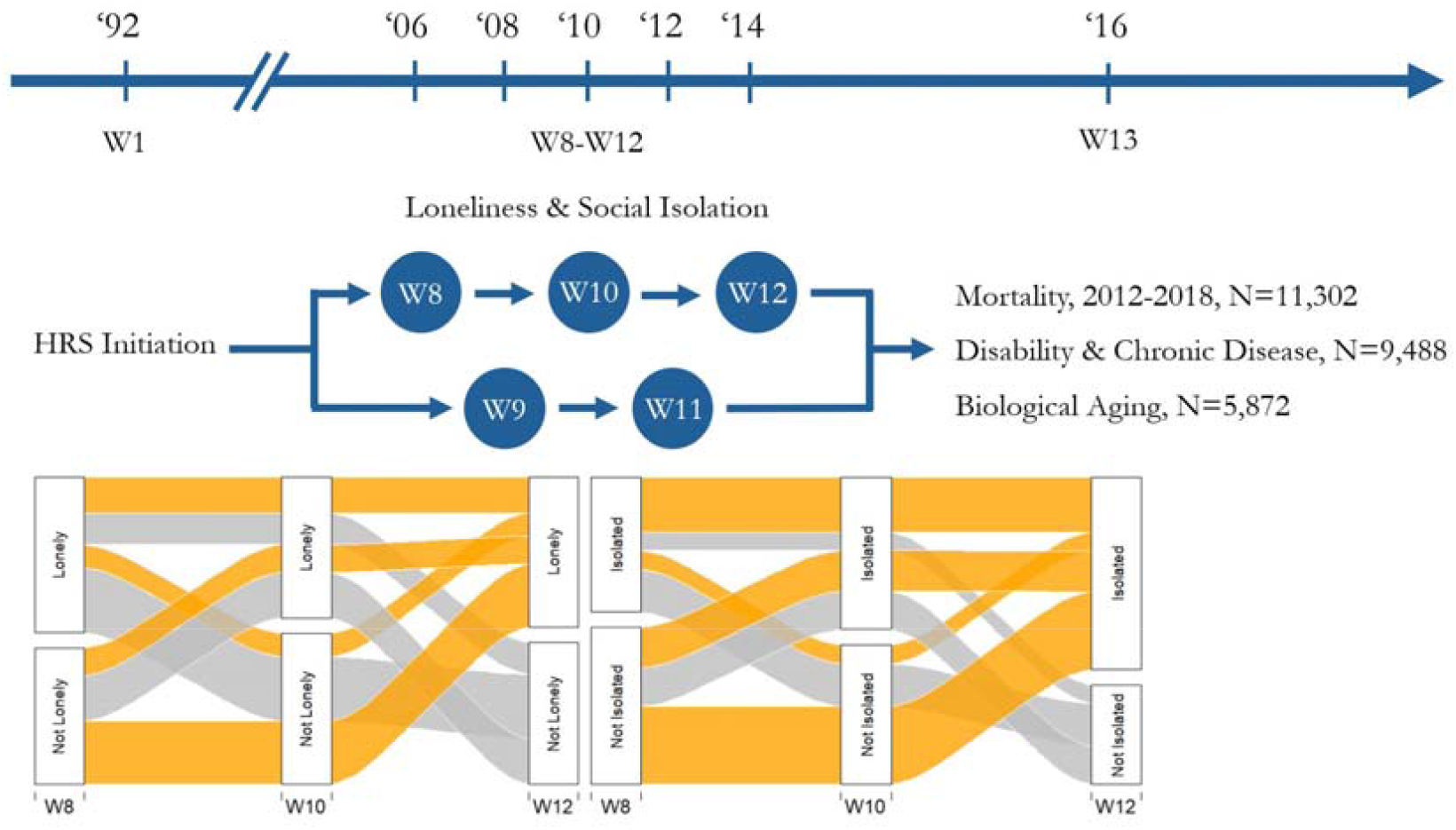
Timeline of assessments of loneliness, social isolation, and deficits in healthy aging. HRS collected loneliness and social isolation data at every other assessment wave with half of the sample first surveyed in Wave 8 (2006) and the other half first surveyed in Wave 9 (2008). We included participants who completed at least two assessments of loneliness and social isolation. We classified participants who met criteria at one wave of measurement as “Intermittent” cases and those who met criteria at two or more waves of measurement as “Persistent” cases. The river plots show trajectories of loneliness and social isolation for participants who were measured at three timepoints and met criteria for loneliness or social isolation at least once during follow-up (N=737 for loneliness; N=961 for social isolation). The thickness of each path is indicative of the proportion of participants that followed each trajectory. For mortality, we analyzed data between the last assessment of loneliness and social isolation (Wave 11 (2012) or Wave 12 (2014)) and Wave 13 (2016). HRS collected data on death such that data recorded in Wave 13 included deaths through 2018. For analysis of disability and chronic disease, we considered prevalent and incident reports of Activities of Daily Living (ADL) or Instrumental Activities of Daily Living (IADL) limitations and chronic disease diagnoses. For analysis of prevalent disability and disease, we used the total number of ADL or IADL limitations and chronic disease diagnoses in 2016. For analysis of incident disability and disease, we used the number of new cases of ADL or IADL limitations and chronic disease diagnoses between participants’ second assessment of loneliness and social isolation and 2016. In the incident analysis, exposure classification was based only on the first two assessments of loneliness and social isolation.

### Statistical Analysis

Our analysis sample included participants aged 50-95 at their baseline observation for experiences of loneliness and exposure to social isolation who provided at least two timepoints of data for these measures during follow-ups from 2006-2014 and were alive at the end of the exposure assessment period (2012 or 2014). Disease and disability analysis included the subset of participants who had data for disease and disability outcomes in 2016, and biological aging analysis included the subset of participants included in the 2016 Venous Blood Study. A comparison of participants included in these samples is reported in **Supplemental Tables 1 and 2**.

We tested associations of loneliness and social isolation with mortality, disability, chronic disease, and biological aging using regression methods. We analyzed time-to-event data on mortality using Cox proportional hazards regression to estimate hazard ratios (HR). Proportional hazards assumptions were met. We analyzed count data on number of ADL and IADL disabilities and chronic disease diagnoses using negative binomial regression models to estimate incidence rate ratios (IRRs). We analyzed continuously distributed data on Phenotypic Age Advancement using linear regression to estimate standardized effect-sizes (Cohen’s *d*). All models included covariate adjustment for age, age-squared, sex, age-sex interactions, race/ethnicity, and a dummy variable coding whether participants were assigned to the subsample of the HRS which first measured loneliness and social isolation in 2006 or 2008. Analysis was performed using Stata 15 (46).

## RESULTS

Analysis included 11,302 participants with at least two repeated measures of loneliness or social isolation who were aged 50-95 when they completed their first psychosocial questionnaire and were alive at the end of the exposure assessment period (2012 or 2014). Sample characteristics are reported in **Supplemental Table 2**. At the first waves of measurement (2006 and 2008) 10% of the sample met criteria for experiences of loneliness and 11% met criteria for exposure to social isolation. By the end of exposure assessment in 2014, the proportion that ever met criteria was 18% for loneliness and 21% for social isolation. Of those who ever met criteria for loneliness or social isolation, 20% ever met criteria for both loneliness and social isolation (**Supplemental Table 3**).

Ever reporting experiences of loneliness or exposure to social isolation during follow-up was more common in women as compared to men (for loneliness, Risk Ratio (RR)=1.34, 95% CI [1.23-1.46]; for social isolation, RR=1.09, [1.02-1.19]). Older participants were less likely than younger participants to report loneliness but more likely to report social isolation (a 10-year increase in age was associated with RR=0.88, [0.84-0.92] for loneliness and RR=1.22, [1.17-1.26] for social isolation). Similarly, white participants were less likely than non-white participants to report loneliness but more likely to report social isolation (loneliness RR=0.76, [0.70-0.83]; social isolation RR=1.07, [0.98-1.16]). These demographic factors were included as covariates in all analyses.

During follow-up, HRS recorded deaths for 1,096 participants in our analysis sample. Participants who ever reported experiences of loneliness or exposure to social isolation during 2006-2014 were at increased risk of mortality compared to those who never reported loneliness or social isolation (for loneliness, Hazard Ratio (HR)=1.48, 95% CI [1.28-1.72]; for social isolation, HR=1.38, [1.21-1.58]).

In 2016, 19% of participants (N=1,763) reported one or more ADL disabilities, 18% (N=1,697) reported one or more IADL disabilities, and 82% (N=7,811) reported one or more chronic diseases. Those who ever reported experiences of loneliness or exposure to social isolation during 2006-2014 reported higher levels of all three outcomes as compared to those who never reported loneliness or social isolation (for loneliness, prevalent-ADL-Incidence Rate Ratio (IRR)=2.01, 95% CI [1.80-2.24], prevalent-IADL-IRR=1.99, [1.78-2.23], prevalent-chronic disease-IRR=1.17, [1.14-1.21]; for social isolation, prevalent-ADL-IRR=1.63, [1.46-1.82], prevalent-IADL-IRR=1.51, [1.35-1.69], prevalent-chronic disease-IRR=1.11, [1.08-1.14]).

**Figure 2** graphs survival proportions and percentages of participants with any disability and multimorbidity by strata of loneliness and social isolation. Effect-sizes and confidence intervals are reported in **Supplemental Table 4**.

**Figure 2.**
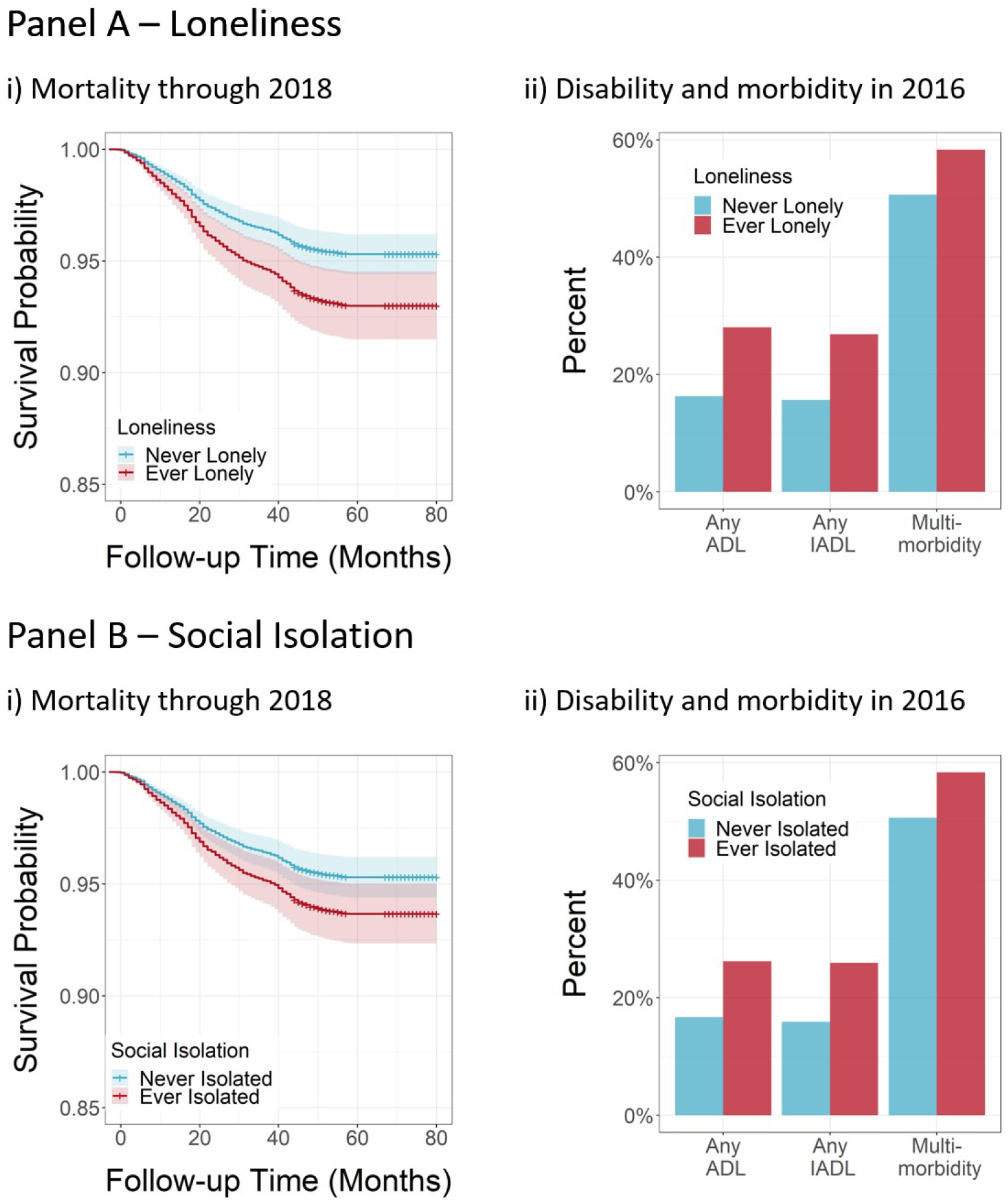
Associations of loneliness and social isolation with deficits in healthy aging. Panels A and B show results from analysis of loneliness and social isolation, respectively. Cell i) plots survival curves for participants who ever reported loneliness or social isolation (red line) and participants who never reported loneliness or social isolation (blue line) estimated from a Cox model including covariate adjustment for age, age-squared, sex, age-sex interactions, race/ethnicity, and a dummy variable coding whether participants were assigned to the subsample of the HRS which first measured loneliness and social isolation in 2006 or 2008. Shaded areas show 95% confidence intervals. Cell ii) plots the percent of participants reporting any Activities of Daily Living (ADL) limitations, any Instrumental Activities of Daily Living (IADL) limitations, and multimorbidity (i.e., two or more chronic disease diagnoses) among participants who ever reported loneliness or social isolation (red bars) and participants who never reported loneliness or social isolation (blue bars).

### Disentangling effects of loneliness and social isolation on deficits in healthy aging from the effects of correlated risk factors

Experiences of loneliness and exposure to social isolation are not randomly distributed throughout the population. Poorer social and economic circumstances and psychological vulnerabilities may put individuals at greater risk for loneliness and social isolation and increase risk for deficits in healthy aging. We therefore repeated our analysis adding covariate adjustment to account for these correlated risk factors. This analysis evaluated confounding of associations between social connectedness and deficits in healthy aging by risk factors present from earlier in life.

We measured participants’ social and economic circumstances from their reports about neighborhood social cohesion and physical disorder, household wealth, and educational attainment. Those from poorer social and economic circumstances more often reported loneliness and social isolation (**Supplemental Table 5**). Social and economic circumstances accounted for some but not all of the associations of loneliness and social isolation with deficits in healthy aging. Covariate adjustment for social and economic circumstances attenuated associations of loneliness and social isolation with all outcomes by 25-37% and 27-53%, respectively (**Figure 3**, green bars and **Supplemental Table 4**).

**Figure 3.**
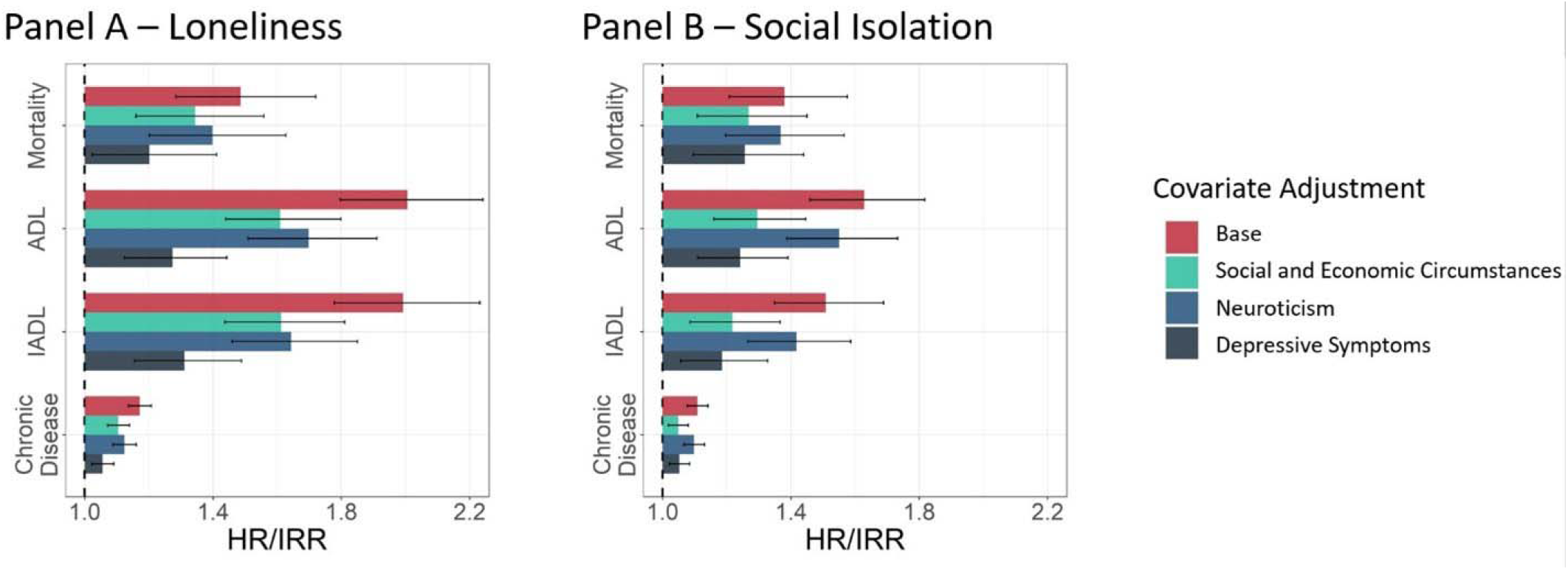
Effect-sizes for associations of loneliness and social isolation with deficits in healthy aging, with adjustment for social and economic circumstances and psychological vulnerabilities. Panels A and B show results from analysis of loneliness and social isolation, respectively, across different covariate adjusted models. The base model included covariate adjustment for age, age-squared, sex, age-sex interactions, race/ethnicity, and a dummy variable coding whether participants were assigned to the subsample of the HRS which first measured loneliness and social isolation in 2006 or 2008. The additional models included these covariates as well as a composite score for social and economic circumstances, a measure of the personality trait neuroticism, and a depressive symptom score. Social and economic circumstances were measured from longitudinal data across all waves of loneliness/social isolation assessment. Neuroticism was measured at the time of the first loneliness/social isolation assessment. Depressive symptoms were measured from 1994-the time of the first loneliness/social isolation assessment. Plots show effect-sizes for analysis of mortality (hazard ratios (HR)) and disability and chronic disease (incidence rate ratios (IRR)), comparing those who ever reported loneliness or social isolation to those who never reported loneliness or social isolation. Error bars represent 95% confidence intervals.

We measured psychological vulnerabilities from baseline reports of the personality trait neuroticism and depressive symptoms. Participants with more psychological vulnerabilities at baseline more often reported loneliness and social isolation (**Supplemental Table 5**). Psychological vulnerabilities accounted for some but not all of the associations of loneliness and social isolation with deficits in healthy aging. Covariate adjustment for baseline levels of neuroticism attenuated associations of loneliness and social isolation with all outcomes by 15-28% and 3-15%, respectively. Covariate adjustment for baseline depressive symptoms attenuated associations of loneliness and social isolation with all outcomes by 54-66% and 29-59%, respectively (**Figure 3**, blue and dark blue bars and **Supplemental Table 4**).

As a further analysis to address confounding by factors that could influence both social connectedness and healthy aging but that were not measured by the HRS, we used fixed-effects regression to test associations of changes in loneliness and social isolation with changes in deficits in healthy aging. Fixed-effects analysis blocks confounding by time-invariant characteristics of individuals that may contribute to both loneliness and social isolation and deficits in healthy aging. We analyzed three deficits in healthy aging with repeated measures in the HRS: ADLs, IADLs, and chronic diseases. Effect-sizes were similar to effect-sizes from covariate adjusted regression models (**Supplemental Table 6**).

### Testing loneliness and social isolation as precursors to deficits in healthy aging

We measured experiences of loneliness and exposure to social isolation prior to the assessment of deficits in healthy aging. However, this prospective design does not rule out the possibility that prior disability and disease might cause loneliness and social isolation. To refine our inference, we limited our measurement of loneliness and social isolation to the first two assessments and conducted analysis of incident disability and chronic disease during the interval between the second assessment of loneliness and social isolation and follow-up in 2016. We included all participants from the main analysis, regardless of whether they were free of any disability or disease at baseline. During follow-up of 2-6 years, 13% of participants (N=1,219) reported one or more incident ADL disabilities, 13% (N=1,235) reported one or more incident IADL disabilities, and 24% (N=2,266) reported one or more incident chronic diseases (**Supplemental Figure 1**).

Participants who reported loneliness and social isolation during the first two psychosocial questionnaires had higher incidence of ADL and IADL disabilities in 2016 (for loneliness, incident-ADL-IRR=1.64, 95% CI [1.41-1.91]; incident-IADL-IRR=1.57, [1.35-1.83]; for social isolation, incident-ADL-IRR=1.35, [1.16-1.57], incident-IADL-IRR=1.32, [1.15-1.53]). Incidence of chronic disease did not differ between participants who reported loneliness and social isolation and those who did not (for loneliness, IRR=1.02, [0.91-1.13]; for social isolation, IRR=0.99, [0.89-1.09]). Effect-sizes are reported in **Supplemental Table 7** and graphed in **Supplemental Figure 1**.

### Comparing intermittent and persistent exposure phenotypes

Of participants who ever met criteria for loneliness during follow-up between 2006-2014, 32% met criteria at multiple assessment waves. We classified these participants as persistently experiencing loneliness (6% of the sample) and the remainder as intermittently experiencing loneliness (12% of the sample). Of participants who ever met criteria for social isolation during follow-up, we classified 37% as persistently exposed to social isolation (8% of the sample) and the remainder as intermittently exposed to social isolation (13% of the sample).

Participants with persistent experiences of loneliness were at increased risk for mortality through 2018 compared to participants with intermittent experiences of loneliness (persistent-loneliness-HR=1.57, 95% CI [1.24-1.99] as compared to intermittent-loneliness-HR=1.45, [1.22-1.72]). In parallel, those with persistent loneliness were at increased risk for prevalent disability and chronic disease (for prevalent ADL disability, persistent-loneliness-IRR=2.57, [2.19-3.02] as compared to intermittent-loneliness-IRR=1.75, [1.53-1.99]; for prevalent IADL disability, persistent-loneliness-IRR=2.34, [1.98-2.77] as compared to intermittent-loneliness-IRR=1.83, [1.60-2.10]; for prevalent chronic disease, persistent-loneliness-IRR=1.22, [1.16-1.28] as compared to intermittent-loneliness-IRR=1.15, [1.11-1.19]). Effect-sizes are graphed in **Figure 4** and reported in **Supplemental Table 8**.

**Figure 4.**
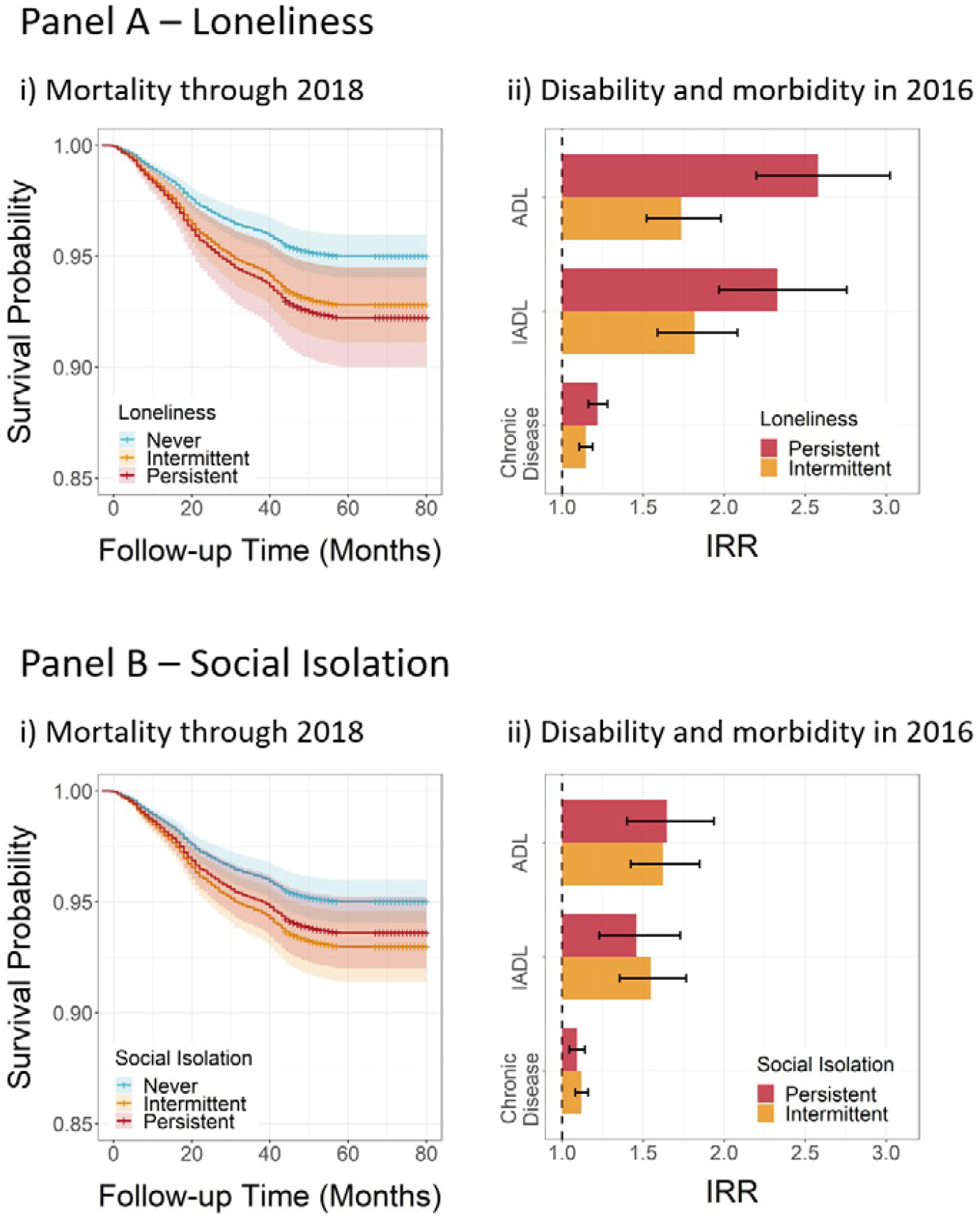
Associations of intermittent and persistent loneliness and social isolation with deficits in healthy aging. Panels A and B show results from analysis of loneliness and social isolation, respectively. Cell i) plots Kaplan Meier survival curves for participants who reported persistent loneliness or social isolation (red line), participants who reported intermittent loneliness or social isolation (orange line), and participants who never reported loneliness or social isolation (blue line). Shaded areas show 95% confidence intervals. Cell ii) plots effect-sizes for analysis of incident Activities of Daily Living disability (ADL), Instrumental Activities of Daily Living disability (IADL), and chronic disease (incidence rate ratios (IRR)), comparing those who reported persistent loneliness or social isolation to those who never reported loneliness or social isolation (red bars) and those who reported intermittent loneliness or social isolation to those who never reported loneliness or social isolation (orange bars). Error bars represent 95% confidence intervals.

In contrast to results for loneliness, we did not find evidence that participants who were persistently exposed to social isolation were at greater risk for any deficits in healthy aging as compared to participants who were intermittently exposed to social isolation (for mortality, persistent-isolation-HR=1.28, 95% CI [1.04-1.56] as compared to intermittent-isolation-HR=1.44, [1.23-1.68]; for prevalent ADL disability, persistent-isolation-IRR=1.65, [1.40-1.94] as compared to intermittent-isolation-IRR=1.62, [1.42-1.85]; for prevalent IADL disability, persistent-isolation-IRR=1.46, [1.23-1.73] as compared to intermittent-isolation-IRR=1.54, [1.35-1.76]; for prevalent chronic disease, persistent-isolation-IRR=1.09, [1.04-1.14] as compared to intermittent-isolation-IRR=1.12, [1.08-1.16]). Effect-sizes are graphed in **Figure 4** and reported in **Supplemental Table 8**.

### Evaluating biological aging as a potential mechanism linking loneliness and social isolation to deficits in healthy aging

We measured participants’ biological aging using the Phenotypic Age algorithm (41–43). As reported previously (47), participants’ Phenotypic Ages were highly correlated with their chronological ages (r=0.76). In our analysis sample (N=5,872), participants’ Phenotypic Ages were, on average, 0.50 years (SD=8.54) older than their chronological ages, indicating that participants’ aging was similar to the expectation based on the NHANES reference sample in which the Phenotypic Age algorithm was developed. Participants who reported more experiences of loneliness exhibited more advanced biological aging (persistent-loneliness-d=0.26, 95% CI [0.14-0.39] as compared to intermittent-loneliness-d=0.12, [0.04-0.20]). For social isolation, participants with any exposure tended to have more advanced biological aging as compared to those never exposed, but there was no evidence of increased risk due to persistent exposure (persistent-isolation-d=0.21, [0.11-0.31] as compared to intermittent-isolation-d=0.19, [0.10-0.27]). The relationships between chronological age and phenotypic age and plots of average Phenotypic Age Advancement across strata of persistence are shown in **Figure 5**. Effect-sizes are reported in **Supplemental Table 8**.

**Figure 5.**
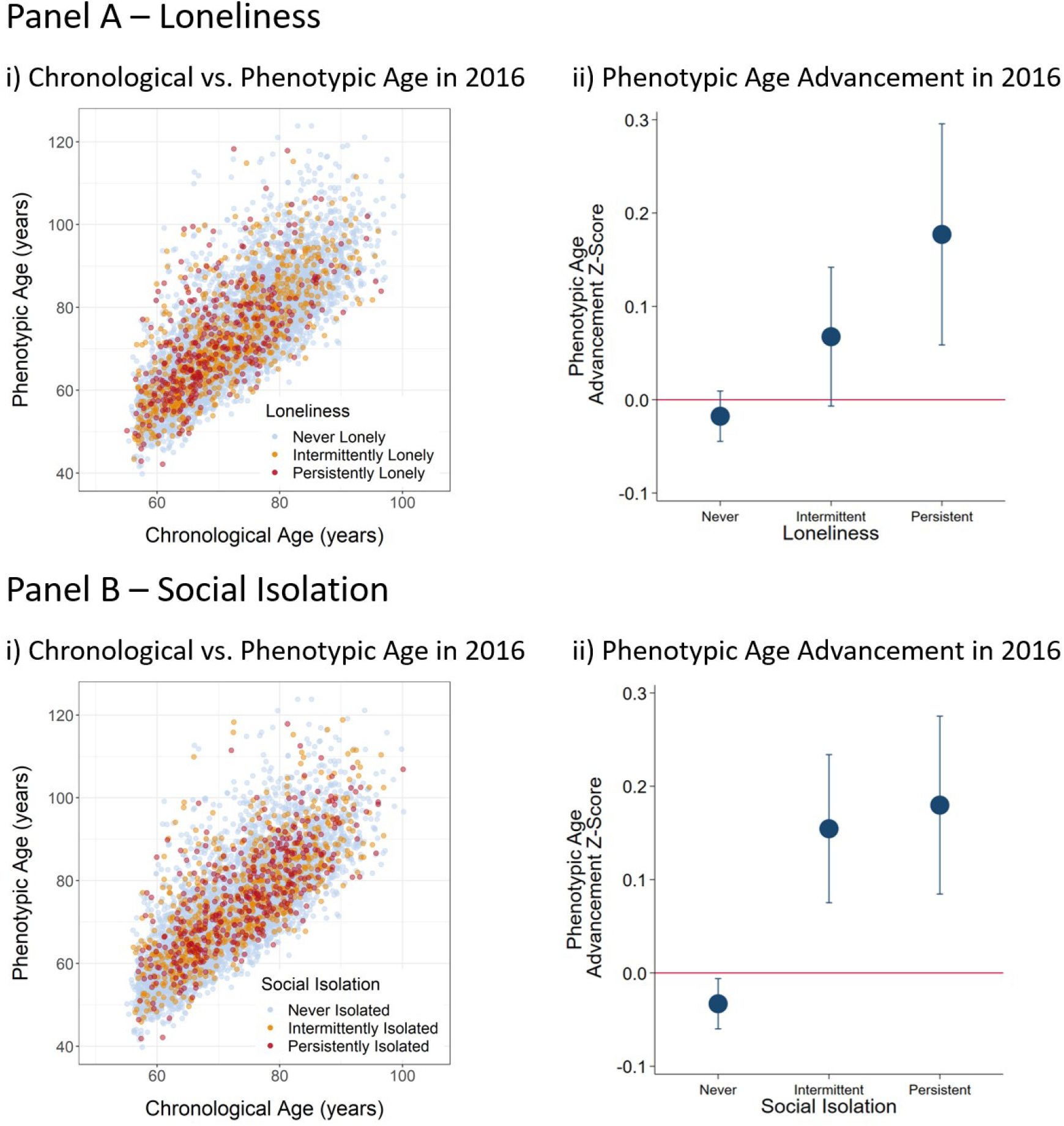
Associations of loneliness and social isolation with biological aging, by levels of loneliness and social isolation persistence. Panels A and B show results from analysis of loneliness and social isolation, respectively. Cell i) shows a scatter plot of chronological age versus Phenotypic Age for participants who reported persistent loneliness or social isolation (red), participants who reported intermittent loneliness or social isolation (orange), and participants who never reported loneliness or social isolation (blue). Cell ii) shows mean Phenotypic Age Advancement (Phenotypic Age – chronological age) for participants exposed to loneliness and social isolation across the strata of Never, Intermittent, and Persistent loneliness and isolation. Phenotypic Age Advancement values are plotted as z-scores (M=0, SD=1). Error bars show 95% confidence intervals.

### Sensitivity Analysis

We conducted sensitivity analyses. First, to test if our findings depended on our measures of experiences of loneliness and exposure to social isolation, we repeated analysis using alternative codings of the 3-item R-UCLA Loneliness Scale and the 6-item social isolation scale as well as alternative measures of social isolation. We also repeated analysis using continuous loneliness and social isolation scores. The results from this sensitivity analysis were generally the same as the results from the main analysis. Results are reported in **Supplemental Table 7** and **Supplemental Figure 2**.

Second, because our analysis sample included a large chronological age range, we compared findings in a younger subset of the sample (age 50-64) to the older subset of the sample (age 65-95). Results were similar in both groups although effect-sizes were somewhat larger for the younger subset. Results are reported in **Supplemental Table 9**.

Third, the group of participants identified as having intermittent experiences of loneliness and exposure to social isolation varied in the timing of their experiences and exposures relative to outcome assessment. Among those who were intermittently lonely or socially isolated, we compared findings for those who were last lonely or socially isolated at their most recent assessment wave and those who were last lonely or socially isolated at earlier assessment waves. Results are reported in **Supplemental Table 10**.

## DISCUSSION

We tested how older adults’ experiences of loneliness and exposure to social isolation were related to deficits in healthy aging using longitudinal, repeated measures data from the HRS. We measured loneliness and social isolation during 2006-2014 and analyzed health outcomes in 2016 and mortality through 2018. Findings add to knowledge about relationships of loneliness and social isolation with deficits in healthy aging in four ways:

First, experiences of loneliness and exposure to social isolation are associated with deficits in healthy aging, and these associations are partly but not fully explained by correlated social and economic circumstances and psychological vulnerabilities that make loneliness and social isolation more likely. This result points to the centrality of social and economic circumstances to healthy aging. It also highlights the challenge of disentangling loneliness and social isolation from mental health symptoms that may be both causes and consequences of deficits in social connectedness. Second, analysis of incident disability and chronic disease ruled out reverse causation as an explanation for the associations of loneliness and social isolation with disability but not in the case of chronic disease. Third, older adults with persistent experiences of loneliness suffered more severe deficits in healthy aging as compared to those with intermittent experiences of loneliness. In contrast, we found no evidence for a similar increased risk due to persistence in the case of exposure to social isolation. Fourth, associations of loneliness and social isolation with deficits in healthy aging were related to an overall process of biological aging. Previous studies have linked loneliness and social isolation with dysregulation of the immune system (30), and our findings suggest that the biology of the relationships of loneliness and social isolation with deficits in healthy aging may encompass quantifiable declines across multiple physiological systems.

These findings must be interpreted within the context of limitations. The measures of experiences of loneliness and exposure to social isolation used in our analysis are imprecise and are not parallel in what they capture. There are no current gold standard measures for the constructs we studied. Misclassification is possible. We used measurements validated within the HRS and its sister-study ELSA, the 3-Item Revised UCLA Loneliness scale (35) and the 6-Item Social Isolation scale (5,36,39). Sensitivity analysis using alternative measures of loneliness and social isolation yielded results similar to those reported in the main analysis (see **Supplemental Methods, Supplemental Table 7, and Supplemental Figure 2**). Loneliness and social isolation were first assessed in the HRS in 2006 and assessment occurred at every other measurement wave. Most participants had only two or three repeated measures. Classification of persistence may change with additional follow-up. In parallel, there is no gold standard measure of aging.

We analyzed deficits in healthy aging using a combination of mortality records, self-reported disability and chronic disease diagnosis data, and a clinical-biomarker-assessed measure of biological aging. Consistent findings across these outcomes bolster confidence in our conclusions. Follow-up time was limited. We were only able to analyze biological aging at a single time point. For analysis of incident disability and disease, prospective follow-up extended at most six years. Continued waves of HRS follow-up will allow for repeated-measures analysis of biological aging and longer follow-up of incident disability and disease outcomes.

Experiences of loneliness and exposure to social isolation may be culturally dependent. Our study was based in the United States, and findings may not be transportable to other settings around the world. Strengths of our study include a longitudinal repeated-measures design, analysis of a large national sample of older adults with measurements of loneliness, social isolation, multiple healthy aging endpoints, and key confounding and mediating factors.

Within the context of these limitations, our findings have implications for research related to loneliness, social isolation, and healthy aging and potentially for public health practice. For research, our findings have three implications. First, better understanding is needed about how and for whom exposure to social isolation results in experiences of loneliness. In alignment with previous research (5,48), not all individuals in the HRS who reported exposure to social isolation also reported experiences of loneliness. An identification of unique types and characteristics of social relationships that link social isolation to loneliness may inform future interventions and allow for more targeted efforts. Additionally, current measures of loneliness and social isolation are crude, and improved measures may better capture the relationship between those who are isolated and those who are lonely. Second, our findings highlight overlap between the effects of histories of depressive symptoms and the effects of loneliness and social isolation. Future studies should build upon previous efforts to investigate the shared etiology of depression and loneliness, for example, through analysis of the shared genetic basis for these conditions (49). Our findings also highlight continued need for longitudinal repeated-measures studies to disentangle the reciprocal nature of causation between depression and loneliness (17,50). Third, the observation that associations of loneliness and social isolation with mortality, disability, and morbidity were also reflected in an advanced state of biological aging suggests the possibility that methods to quantify biological aging, such as the Phenotypic Age algorithm used in this study, may provide sensitive endpoints for intervention trials. In our study, disease incidence over up to six years was unrelated to loneliness or social isolation. Thus, timescales for most intervention follow-up may not be sufficient to detect impact on disease risk. Because methods to quantify biological aging focus on changes that precede disease onset, they may be more sensitive to near-term biological changes resulting from enhanced social connectedness.

For public health practice, our findings amplify prior work identifying experiences of loneliness as the proximate determinant of deficits in healthy aging. Proposed interventions aim to improve health outcomes by reducing the length of experiences of loneliness and exposure to social isolation (13). In our analysis, a less-persistent phenotype was associated with reduced risk only in the case of loneliness. Deficits in healthy aging associated with social isolation were similar across levels of persistence, raising the possibility that interventions reducing length of exposure to social isolation without directly impacting experiences of loneliness may not improve health outcomes.

Our overall findings support a relationship of experiences of loneliness and exposure to social isolation with deficits in healthy aging and provide further motivation for intervention trials. They nevertheless highlight two enduring challenges facing research to understand the public health impacts of loneliness and social isolation and efforts to design effective interventions: First, deficits in healthy aging are more concentrated in those individuals who experience persistent deficits in social connectedness. But these individuals represent a minority of the overall population experiencing loneliness or being exposed to social isolation at any given point in time. Longitudinal phenotyping will be important for advancing understanding of etiology and impact. Second, liability to experiences of loneliness and exposure to social isolation is variable in the population and risk is greater in those with few socioeconomic resources and who struggle with mental health problems. Tailoring interventions to meet the needs of these vulnerable populations will be critical.

## Data Availability

All data are publicly available on the Health and Retirement Study (HRS) website.

## Author Contributions

The study was designed by DWB and CLC. CLC conducted all analyses with supervision from DWB and support from GH and DK. CLC and DWB wrote the paper. All authors contributed critical feedback related to study design and execution and to manuscript preparation and revision.

We are grateful to Linda P. Fried and to the Psychiatric Epidemiology Training Program at the Columbia University Mailman School of Public Health for feedback on earlier drafts of this manuscript.

## SUPPLEMENTAL MATERIALS

### SUPPLEMENTAL METHODS

#### Covariate Descriptions

We used data from RAND corporation and the Leave Behind Questionnaire (LBQ) to gather information on other variables of interest. We extracted data on gender, race/ethnicity, household wealth, educational attainment, and depression from the RAND HRS Longitudinal File includes. We extracted data on neighborhood social cohesion, neighborhood physical disorder, and neuroticism from the LBQ.

#### Demographic Covariates

RAND considered gender a binary variable, either male or female. We derived race/ethnicity from one question regarding race and another question regarding Hispanic/Latino heritage.

#### Social and Economic Circumstances

We measured participants’ social and economic circumstances across three domains: neighborhood conditions, household wealth, and education.

We measured neighborhood conditions from reports about neighborhood social cohesion and physical disorder (scored using two 4-item scales (1)(44)). The neighborhood social cohesion index is based on the extent to which participants felt people in their neighborhood: 1) make them feel like they do not belong, 2) cannot be trusted, 3) are not friendly, and 4) are not helpful in times of need. The physical disorder index is based on the extent to which participants felt their neighborhood: 1) has a problem with vandalism, 2) is an unsafe place to walk alone after dark, 3) is dirty, and 4) has many vacant buildings. Participants responded to each of these eight items using a seven-point scale. We reverse coded responses so that higher scores indicate more social cohesion and less physical disorder. For both measures, we calculated a score for each wave by averaging the responses for those who responded to at least two of the four items. Finally, we calculated a z-score based on the average scores for participants from 2006 to 2014.

We measured household wealth from dollar values computed by RAND corporation based on structured interviews with participants about their assets. Following the method we used previously (2), values were inflated to constant dollars, inverse-hyperbolic-sign transformed to reduce skew, standardized by age and sex, and averaged across measurement waves, resulting in a single wealth value for each participant.

We measured education as the highest level of educational attainment (coded 1-3 for less than high school, completed high school, and completed college or more).

We calculated a composite score for socioeconomic circumstances as the sum of z-scores across neighborhood, wealth, and education domains.

#### Psychological Vulnerabilities

We measured participants’ psychological vulnerabilities in two ways.

First, we measured the personality trait neuroticism. Neuroticism was assessed with a 4-item scale (3) at the time of the participant’s first assessment of loneliness and social isolation. Participants reported the extent to which they would use the following words to describe themselves: 1) moody, 2) worrying, 3) nervous, and 4) calm. We reverse coded all items except “calm” so higher scores indicate higher levels of neuroticism.

Second, we measured depressive symptoms. The HRS measured participants’ depressive symptoms using the 8-item Center for Epidemiologic Studies Depression Scale (CES-D, (4, 5)) at all waves from 1994 on. However, given that one of the independent variables in this study is loneliness, we removed one item on loneliness and the depression score was based on how frequently in the past week the respondent felt: 1) depressed, 2) everything was an effort, 3) sleep was restless, 4) happy, 5) life was enjoyable, 6) sad, unable to get going. We coded responses so higher scores indicate more severe depressive symptoms. Others have followed this method to avoid item overlap and have found minimal decreases in internal consistency of the scale (6-8). To measure long-term risk for depression, we averaged scores from all waves until the participant’s first assessment of loneliness and social isolation.

#### Sensitivity Analysis

In our main analysis, we classified participants as lonely if their scale scores exceeded the 80^th^ percentile of the cohort distribution (scores ≥7 and ≥3 for loneliness and social isolation, respectively), following the procedure of Steptoe et al. (9). We conducted sensitivity analysis to test if our findings depended on our measures of loneliness and social isolation.

#### Loneliness

For loneliness, we repeated our analysis with two less restrictive classifications of the 3-item Revised UCLA (R-UCLA) Loneliness Scale. First, we used a cut-point published for the English Longitudinal Study of Aging (ELSA), classifying participants as lonely if they had scores ≥6 (9). Second, we used a cut-point published for the HRS, classifying participants as lonely if they responded “some of the time” to any of three items asking about how frequently participants felt they were lacking companionship, left out, and isolated from others (10). In addition to these alternative classifications, we also analyzed the 3-item UCLA loneliness scale scores as a continuous measure. We calculated averages for each participant’s loneliness score from 2006-2014 and then computed z-scores based on these averages.

#### Social Isolation

For social isolation, we repeated our analysis with a less restrictive classification of the 6-item social isolation scale and two alternative measures. First, we used the 6-item social isolation scale with a cut-point published for the ELSA (scores ≥2) (9). Second, we used a 10-item social isolation scale originally developed for the National Social Life, Health, and Aging Project (NSHAP) (11). After being adapted to the HRS, it included 10 items: items 1-3 counted the number of children (coded 0-5+), other family members (coded 0-10+), and friends (coded 0-10+). Items 4-6 counted the frequency of contact with each of these relationships (coded 1-6 for the responses: 1) never/<1 time per year, 2) 1/2 times per year, 3) every few months, 4) 1/2 times per month, 5) 1/2 times per week, and 6) 3+ times per week). Item 7 counted church attendance (coded 1-5 for the responses: 1) never, 2) 1+ times per year, 3) 2/3 times per month, 4) 1 time per week, and 5) 1+ times per week). Item 8 counted other organization/group attendance (coded 1-6 for the responses: 1) never, 2) <1 time per month, 3) about 1 time per month, 4) several times per month, 5) 1 time per week, and 6) 1+ times per week). Item 9 indicated whether or not a participant volunteered in the past year (coded 0 or 1). Item 10 counted the number of people living with the participant (coded 0-5+) (8). For each item, we computed z-scores based on the distribution of values in the baseline interview (2006 or 2008 depending on which wave participants were administered their first psychosocial questionnaire). We then averaged z-scores across items to compute the 10-item social isolation scale for those who responded to at least 5 of the 10 items. Participants who scored 1 SD above the mean or higher were classified as socially isolated. Third, we used a 4-item scale developed for the National health and Aging Trends Study (NHATS) (12), which we adapted to the HRS. After being adapted to the HRS, participants received one point for each of the following: 1) not living alone; 2) being able to open up “a lot” to their spouse, children, other family members, or friends; 3) attending religious services 2 or 3 times a month; and 4) participating in community organizations at least once a month. We created a sum score for those who responded to at least 2 of the items. Following the methods used by Cudjoe et al. (12), we classified participants as socially isolated if their scale scores were ≤1. In addition to these alternative classifications, we also analyzed the 6-item social isolation scale scores as a continuous measure. We calculated averages for each participant’s social isolation score from 2006-2014 and then computed z-scores based on these averages.

## SUPPLEMENTAL TABLES

**Supplemental Table 1.** Sample Characteristics. The table shows summary statistics for age at baseline, sex, ethnicity, social and economic circumstances, and depressive symptoms for participants who received at least one Leave Behind Questionnaire (LBQ), participants who completed at least one LBQ, and participants who were included in the analysis sample. The analysis sample was restricted to those who had repeated-measures data (i.e., two or more measures) on loneliness or social isolation and were aged 50-95 when they completed their first LBQ.

|  | Received 1+ LBQ | Completed 1+ LBQ | Analysis Sample |
| --- | --- | --- | --- |
| **Baseline Age in Years** | **N = 24,890** | **N = 20,674** | **N = 11,302** |
| Mean | 56.96 | 56.92 | 56.31 |
| **Sex** | **N = 24,890** | **N = 20,674** | **N=11,302** |
| % Male | 42 | 41 | 40 |
| % Female | 58 | 59 | 60 |
| **Ethnicity** | **N = 24,879** | **N = 20,667** | **N=11,302** |
| % White, non-Hispanic | 65 | 69 | 75 |
| % Black, non-Hispanic | 19 | 17 | 14 |
| % Latino/Hispanic | 13 | 11 | 9 |
| % Other, non-Hispanic | 3 | 3 | 2 |
| **Social and Economic Circumstances** | **N =24,890** | **N =20,674** | **N = 11,302** |
| Mean | 0.01 | 0.05 | 0.12 |
| **Depressive Symptoms** | -- | **N = 20,527** | **N = 11,251** |
| Mean | -- | 0.00 | -0.09 |

**Supplemental Table 2.**
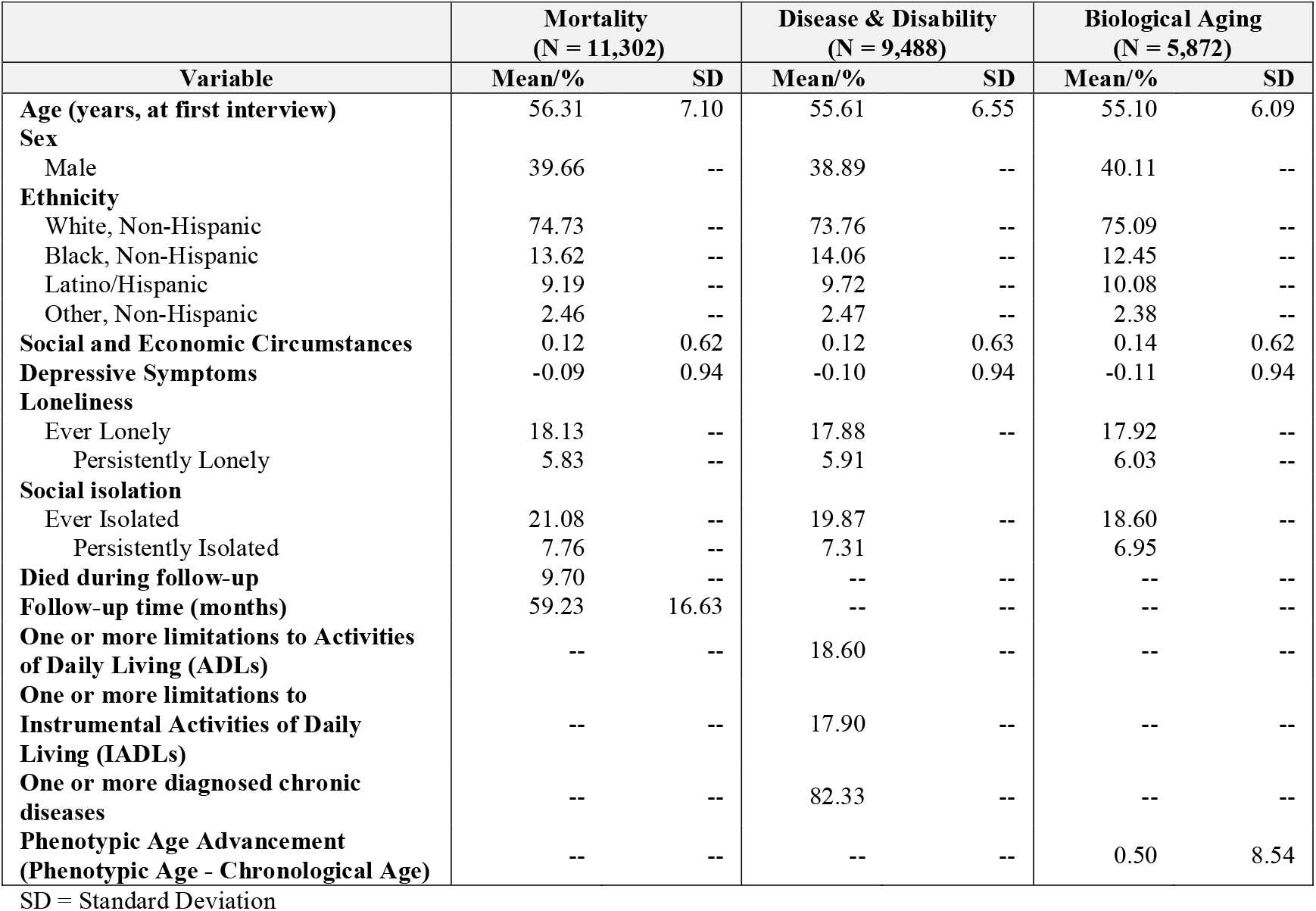
Summary statistics. The table shows summary statistics for demographic, exposure, and outcome variables for participants included in the analysis samples for mortality, disease and disability, and biological aging.

|  | **Mortality**<br>**(N = 11,302)** |  | **Disease & Disability**<br>**(N = 9,488)** |  | **Biological Aging**<br>**(N = 5,872)** |  |
| --- | --- | --- | --- | --- | --- | --- |
| **Variable** | **Mean/%** | **SD** | **Mean/%** | **SD** | **Mean/%** | **SD** |
| **Age (years, at first interview)** | 56.31 | 7.10 | 55.61 | 6.55 | 55.10 | 6.09 |
| **Sex** |  |  |  |  |  |  |
| Male | 39.66 | -- | 38.89 | -- | 40.11 | -- |
| **Ethnicity** |  |  |  |  |  |  |
| White, Non-Hispanic | 74.73 | -- | 73.76 | -- | 75.09 | -- |
| Black, Non-Hispanic | 13.62 | -- | 14.06 | -- | 12.45 | -- |
| Latino/Hispanic | 9.19 | -- | 9.72 | -- | 10.08 | -- |
| Other, Non-Hispanic | 2.46 | -- | 2.47 | -- | 2.38 | -- |
| **Social and Economic Circumstances** | 0.12 | 0.62 | 0.12 | 0.63 | 0.14 | 0.62 |
| **Depressive Symptoms** | -0.09 | 0.94 | -0.10 | 0.94 | -0.11 | 0.94 |
| **Loneliness** |  |  |  |  |  |  |
| Ever Lonely | 18.13 | -- | 17.88 | -- | 17.92 | -- |
| Persistently Lonely | 5.83 | -- | 5.91 | -- | 6.03 | -- |
| **Social isolation** |  |  |  |  |  |  |
| Ever Isolated | 21.08 | -- | 19.87 | -- | 18.60 | -- |
| Persistently Isolated | 7.76 | -- | 7.31 | -- | 6.95 | -- |
| **Died during follow-up** | 9.70 | -- | -- | -- | -- | -- |
| **Follow-up time (months)** | 59.23 | 16.63 | -- | -- | -- | -- |
| **One or more limitations to Activities of Daily Living (ADLs)** | -- | -- | 18.60 | -- | -- | -- |
| **One or more limitations to Instrumental Activities of Daily Living (IADLs)** | -- | -- | 17.90 | -- | -- | -- |
| **One or more diagnosed chronic diseases** | -- | -- | 82.33 | -- | -- | -- |
| **Phenotypic Age Advancement (Phenotypic Age - Chronological Age)** | -- | -- | -- | -- | 0.50 | 8.54 |

SD = Standard Deviation

**Supplemental Table 3.** Co-occurrence of loneliness and social isolation. The table shows a cross-tabulation of ever vs. never meeting criteria for loneliness and ever vs. never meeting criteria for social isolation.

|  | Never Lonely | Ever Lonely |
| --- | --- | --- |
| Never Socially Isolated | 7,058 | 1,182 |
| Ever Socially Isolated | 1,434 | 674 |

**Supplemental Table 4.** Effect-sizes for associations of loneliness and social isolation with deficits in healthy aging before and after covariate adjustment for social and economic circumstances and psychological vulnerabilities. The tables show effect-sizes for analysis of mortality, prevalent Activities of Daily Living (ADL) disability, prevalent Instrumental Activities of Daily Living (IADL) disability, and prevalent chronic disease from five regression models. The first rows report effect-sizes for the base models, which included covariates for age, age-squared, sex, age-sex interactions, race/ethnicity, and a dummy variable coding whether participants were assigned to the subsample of the HRS which first measured loneliness and social isolation in 2006 or 2008. The second through fourth rows report effect-sizes from models adjusted for each of the social and economic circumstances and psychological vulnerability covariates. The final rows report effect-sizes from models that included all of the covariates. The first two columns for each outcome report effect-sizes and 95% confidence intervals (CI). The third column for each outcome reports the percent reduction in the coefficient estimate between the base and the adjusted models. Percent reductions are calculated based on unexponentiated coefficients. For mortality, effect-sizes are reported as hazard ratios (HR). For ADL disability, IADL disability, and chronic disease, effect-sizes are reported as incidence rate ratios (IRR). Panels A and B report analysis of loneliness and social isolation, respectively.

Panel A – Loneliness
|  | Mortality |  |  | Prevalent ADL |  |  | Prevalent IADL |  |  | Prevalent Chronic Disease |  |  |
| --- | --- | --- | --- | --- | --- | --- | --- | --- | --- | --- | --- | --- |
| Model | HR | 95% CI | % Reduction* | IRR | 95% CI | % Reduction | IRR | 95% CI | % Reduction | IRR | 95% CI | % Reduction |
| Base Model | 1.48 | 1.28-1.72 | -- | 2.01 | 1.80-2.24 | -- | 1.99 | 1.78-2.23 | -- | 1.17 | 1.14-1.21 | -- |
| Social and Economic Circumstances | 1.34 | 1.16-1.55 | 25 | 1.61 | 1.44-1.80 | 32 | 1.61 | 1.44-1.81 | 31 | 1.11 | 1.07-1.14 | 37 |
| Neuroticism (baseline) | 1.40 | 1.20-1.62 | 15 | 1.70 | 1.51-1.91 | 24 | 1.64 | 1.46-1.85 | 28 | 1.12 | 1.09-1.16 | 26 |
| 7-Item CES-D ('94-baseline) | 1.20 | 1.02-1.41 | 54 | 1.27 | 1.12-1.44 | 65 | 1.31 | 1.16-1.49 | 61 | 1.06 | 1.02-1.09 | 66 |
| Full Model | 1.16 | 0.98-1.36 | 63 | 1.16 | 1.02-1.32 | 79 | 1.19 | 1.04-1.35 | 75 | 1.03 | 0.99-1.06 | 84 |

Panel B – Social Isolation
|  | Mortality |  |  | Prevalent ADL |  |  | Prevalent IADL |  |  | Prevalent Chronic Disease |  |  |
| --- | --- | --- | --- | --- | --- | --- | --- | --- | --- | --- | --- | --- |
| Model | HR | 95% CI | % Reduction | IRR | 95% CI | % Reduction | IRR | 95% CI | % Reduction | IRR | 95% CI | % Reduction |
| Base Model | 1.38 | 1.20-1.57 | -- | 1.63 | 1.46-1.82 | -- | 1.51 | 1.35-1.69 | -- | 1.11 | 1.08-1.14 | -- |
| Social and Economic Circumstances | 1.26 | 1.10-1.45 | 27 | 1.30 | 1.16-1.45 | 47 | 1.22 | 1.09-1.37 | 52 | 1.05 | 1.02-1.08 | 53 |
| Neuroticism (baseline) | 1.36 | 1.19-1.56 | 3 | 1.55 | 1.39-1.73 | 10 | 1.42 | 1.27-1.59 | 15 | 1.10 | 1.07-1.13 | 9 |
| 7-Item CES-D ('94-baseline) | 1.25 | 1.09-1.44 | 29 | 1.24 | 1.11-1.39 | 56 | 1.18 | 1.06-1.33 | 59 | 1.05 | 1.02-1.08 | 50 |
| Full Model | 1.21 | 1.05-1.39 | 41 | 1.12 | 1.00-1.25 | 77 | 1.07 | 0.96-1.21 | 83 | 1.03 | 0.99-1.06 | 76 |

\*% reduction is calculated based on unexponentiated coefficients

**Supplemental Table 5.** Effect-sizes for associations of social and economic circumstances and psychological vulnerabilities with the number of waves at which participants were classified as lonely or socially isolated. The table reports effect-sizes as incidence rate ratios estimated from negative binomial regressions for associations with the number of waves at which participants were classified as lonely or socially isolated. All models included covariate adjustment for age, age-squared, sex, age-sex interactions, race/ethnicity, and a dummy variable coding whether participants were assigned to the subsample of the HRS which first measured loneliness and social isolation in 2006 or 2008. For neighborhood social cohesion, neighborhood physical disorder, and household wealth, higher scores indicate better social and economic circumstances. For neuroticism and depressive symptoms, higher scores indicate more psychological vulnerabilities.

|  | 3-Item UCLA Loneliness |  | 6-Item Social Isolation |  |
| --- | --- | --- | --- | --- |
|  | IRR | 95% CI | IRR | 95% CI |
| **Neighborhood Social Cohesion** | 0.59 | 0.56-0.61 | 0.74 | 0.71-0.77 |
| **Neighborhood Physical Disorder** | 0.74 | 0.71-0.78 | 0.81 | 0.78-0.85 |
| **Household Wealth** | 0.64 | 0.61-0.66 | 0.58 | 0.56-0.60 |
| **Educational Attainment** |  |  |  |  |
| Less than high school | 1.24 | 1.11-1.39 | 1.19 | 1.07-1.32 |
| Completed high school | Ref | Ref | Ref | Ref |
| Completed college or more | 0.72 | 0.64-0.81 | 0.66 | 0.59-0.74 |
| **Neuroticism (baseline)** | 1.87 | 1.79-1.94 | 1.15 | 1.11-1.20 |
| **Depressive Symptoms (baseline)** | 1.87 | 1.81-1.93 | 1.43 | 1.38-1.48 |

**Supplemental Table 6.** Marginal effects for associations of loneliness and social isolation with deficits in healthy aging. The table reports marginal effects for associations of loneliness (Panel A) and social isolation (Panel B) with deficits in healthy aging (numbers of Activities of Daily Living (ADL) disabilities, Instrumental Activities of Daily Living (IADL) disabilities, and chronic disease diagnoses) from 3 models. Base Model estimates (far left column) are from models adjusted for age (quadratic), sex, age-sex interactions, and race/ethnicity. These estimates correspond to the effect-sizes graphed in red in Figure 3. Adjusted Model estimates (second column) are from models including additional covariates for social and economic circumstances, neuroticism, and depression. These estimates correspond to the effect-sizes reported in the final row of Supplemental Table 4. Fixed-effect Model estimates (far right columns) are from within-person fixed-effects Poisson regression with covariate adjustment for chronological age. Fixed-effects models test if changes in loneliness and social isolation over the four-year measurement interval were associated with changes in deficits in healthy aging over the same measurement interval. Fixed-effects models block confounding by time-invariant characteristics of individuals that may contribute to both predictor and response variables. The table shows (i) that adjusted and fixed-effects model estimates are substantially attenuated relative to estimates from the base model; and (ii) that for associations with chronic disease, fixed-effects model estimates are near zero.

Panel A – Loneliness
| Outcome | Base Model | Adjusted Model | Fixed-Effects Model | Fixed-Effects Model 95% CI |
| --- | --- | --- | --- | --- |
| ADL | 0.17 | 0.03 | 0.03 | 0.00-0.05 |
| IADL | 0.15 | 0.03 | 0.04 | 0.01-0.07 |
| Chronic Disease | 0.26 | 0.04 | 0.01 | 0.00-0.02 |

Panel B – Social Isolation
| Outcome | Base Model | Adjusted Model | Fixed-Effects Model | Fixed-Effects Model 95% CI |
| --- | --- | --- | --- | --- |
| ADL | 0.12 | 0.02 | 0.04 | 0.02-0.07 |
| IADL | 0.10 | 0.01 | 0.06 | 0.03-0.08 |
| Chronic Disease | 0.17 | 0.04 | -0.01 | -0.02-0.00 |

**Supplemental Table 7.** Effect-sizes for associations of loneliness and social isolation with deficits in healthy aging. Panels A and B report analysis of loneliness and social isolation, respectively. The panels report effect-sizes for the measures of loneliness and social isolation reported in the main text (far left column) along with two alternative codings of loneliness, one alternative coding of social isolation, and two alternative measures of social isolation proposed in previous studies. We classified participants as lonely if they (1) scored ≥7 on the 3-item R-UCLA (31), (2) scored ≥6 on the R-UCLA (31), and (3) responded “some of the time” to any of the 3-item R-UCLA (2). We classified participants as socially isolated if they (1) scored ≥3 on the 6-item scale (31), (2) scored ≥2 on the 6-item scale (31), (3) scored 1 standard deviation above the mean on the 10-item scale (45), and (4) scored ≤1 on the 4-item scale (1). Effect-sizes and 95% confidence intervals are reported across measures. For mortality, effect-sizes are reported as hazard ratios (HR). For prevalent and incident Activities of Daily Living (ADL) disability, Instrumental Activities of Daily Living (IADL) disability, and chronic disease, effect-sizes are reported as incidence rate ratios (IRR). Analysis of incident disability and disease included 1,219 participants with at least one new case of ADL disability, 1,235 participants with at least one new case of IADL disability, and 2,266 participants with at least one new chronic disease diagnosis. All models included covariate adjustment for age, age-squared, sex, age-sex interactions, race/ethnicity, and a dummy variable coding whether participants were assigned to the subsample of the HRS which first measured loneliness and social isolation in 2006 or 2008.

Panel A – Loneliness
| | 3-Item UCLA Loneliness<br>(Lonely: $\geq 7$, 18%) | | 3-Item UCLA Loneliness<br>(Lonely: $\geq 6$, 38%) | | 3-Item UCLA Loneliness<br>(Lonely: any “some of the time” response, 73%) | |
| --- | --- | --- | --- | --- | --- | --- |
|  | Effect-Size | 95% CI | Effect-Size | 95% CI | Effect-Size | 95% CI |
|  | *Mortality (HR) (N = 11,095)* |  |  |  |  |  |
| **Mortality** | 1.48 | 1.28-1.72 | 1.47 | 1.30-1.66 | 1.47 | 1.26-1.70 |
|  | *Prevalent Disability and Disease (IRR) (N = 9,340)* |  |  |  |  |  |
| **ADL** | 2.01 | 1.80-2.24 | 1.94 | 1.76-2.14 | 1.95 | 1.71-2.22 |
| **IADL** | 1.99 | 1.78-2.23 | 1.89 | 1.71-2.09 | 1.85 | 1.62-2.11 |
| **Chronic Disease** | 1.17 | 1.14-1.21 | 1.14 | 1.11-1.17 | 1.13 | 1.10-1.17 |
|  | *Incident Disability and Disease (IRR) (N = 9,340)* |  |  |  |  |  |
| **ADL** | 1.64 | 1.41-1.91 | 1.63 | 1.44-1.84 | 1.57 | 1.36-1.82 |
| **IADL** | 1.57 | 1.35-1.83 | 1.54 | 1.36-1.74 | 1.59 | 1.38-1.83 |
| **Chronic Disease** | 1.02 | 0.91-1.13 | 1.03 | 0.95-1.11 | 1.09 | 1.00-1.19 |

Panel B – Social Isolation
| | 6-Item Social Isolation<br>(Isolated: $\geq 3$, 21%) | | 6-Item Social Isolation<br>(Isolated: $\geq 2$, 49%) | | 10-Item Social Isolation<br>(Isolated: 1 SD above mean, 25%) | | 4-Item Social Isolation<br>(Isolated: $\leq 1$, 20%) | |
| --- | --- | --- | --- | --- | --- | --- | --- | --- |
|  | Effect-Size | 95% CI | Effect-Size | 95% CI | Effect-Size | 95% CI | Effect-Size | 95% CI |
|  | *Mortality (HR) (N = 11,275)* |  |  |  |  |  |  |  |
| **Mortality** | 1.38 | 1.21-1.58 | 1.36 | 1.19-1.54 | 1.49 | 1.31-1.70 | 1.55 | 1.36-1.77 |
|  | *Prevalent Disability and Disease (IRR) (N = 9,467)* |  |  |  |  |  |  |  |
| **ADL** | 1.63 | 1.46-1.82 | 1.44 | 1.30-1.59 | 1.57 | 1.40-1.75 | 1.56 | 1.39-1.75 |
| **IADL** | 1.51 | 1.35-1.69 | 1.33 | 1.19-1.47 | 1.47 | 1.32-1.65 | 1.41 | 1.26-1.59 |
| **Chronic Disease** | 1.11 | 1.08-1.14 | 1.09 | 1.06-1.12 | 1.13 | 1.10-1.16 | 1.11 | 1.07-1.14 |
|  | *Incident Disability and Disease (IRR) (N = 9,467)* |  |  |  |  |  |  |  |
| **ADL** | 1.35 | 1.16-1.57 | 1.27 | 1.12-1.43 | 1.33 | 1.15-1.53 | 1.24 | 1.07-1.45 |
| **IADL** | 1.32 | 1.15-1.53 | 1.29 | 1.14-1.45 | 1.31 | 1.14-1.51 | 1.23 | 1.06-1.43 |
| **Chronic Disease** | 0.99 | 0.89-1.09 | 1.08 | 1.00-1.17 | 1.01 | 0.92-1.11 | 1.04 | 0.94-1.15 |

**Supplemental Table 8.** Effect-sizes for associations of loneliness and social isolation (persistent vs. intermittent vs. never) with deficits in healthy aging. The table reports effect-sizes for associations of intermittent or persistent loneliness or social isolation compared to never loneliness or social isolation with deficits in healthy aging. For associations with mortality, effect-sizes are reported as hazard ratios (HR) estimated from Cox proportional hazards models. For prevalent Activities of Daily Living (ADL) disability, Instrumental Activities of Daily Living (IADL) disability, and chronic disease, effect-sizes are reported as incidence rate ratios (IRR) estimated from negative binomial regression models. For Phenotypic Age Advancement, effect-sizes are reported as standardized regression coefficients estimated from linear regression models, interpretable as Cohen’s *d* (d). All models included covariate adjustment for age, age-squared, sex, age-sex interactions, race/ethnicity, and a dummy variable coding whether participants were assigned to the subsample of the HRS which first measured loneliness and social isolation in 2006 or 2008.

|  | 3-Item UCLA Loneliness |  | 6-Item Social Isolation |  |
| --- | --- | --- | --- | --- |
|  | Effect-Size | 95% CI | Effect-Size | 95% CI |
| *Mortality (HR)* |  |  |  |  |
| **Mortality** |  |  |  |  |
| Intermittent | 1.45 | 1.22-1.72 | 1.44 | 1.23-1.68 |
| Persistent | 1.57 | 1.24-1.99 | 1.28 | 1.04-1.56 |
| *Prevalent Disability and Disease (IRR)* |  |  |  |  |
| **ADL** |  |  |  |  |
| Intermittent | 1.75 | 1.53-1.99 | 1.62 | 1.42-1.85 |
| Persistent | 2.57 | 2.19-3.02 | 1.65 | 1.40-1.94 |
| **IADL** |  |  |  |  |
| Intermittent | 1.83 | 1.60-2.10 | 1.54 | 1.35-1.76 |
| Persistent | 2.34 | 1.98-2.77 | 1.46 | 1.23-1.73 |
| **Chronic Disease** |  |  |  |  |
| Intermittent | 1.15 | 1.11-1.19 | 1.12 | 1.08-1.16 |
| Persistent | 1.22 | 1.16-1.28 | 1.09 | 1.04-1.14 |
| *Biological Aging (d)* |  |  |  |  |
| **Phenotypic Age Advancement** |  |  |  |  |
| Intermittent | 0.12 | 0.04-0.20 | 0.19 | 0.10-0.27 |
| Persistent | 0.26 | 0.14-0.39 | 0.21 | 0.11-0.31 |

**Supplemental Table 9.** Association of loneliness and social isolation (ever vs. never) with deficits in healthy aging, stratified by age (50-64 years vs. 65-95 years). Panels A and B reports effect-sizes for associations of ever being lonely or socially isolated, respectively, compared to never being lonely or socially isolated with deficits in healthy aging. Results are stratified by age to compare effect-sizes for those who are 50-64 years old and those who are 65-95 years old. For associations with mortality, effect-sizes are reported as hazard ratios (HR) estimated from Cox proportional hazards models. For prevalent Activities of Daily Living (ADL) disability, Instrumental Activities of Daily Living (IADL) disability, and chronic disease, effect-sizes are reported as incidence rate ratios (IRR) estimated from negative binomial regression models. All models included covariate adjustment for age, age-squared, sex, age-sex interactions, race/ethnicity, and a dummy variable coding whether participants were assigned to the subsample of the HRS which first measured loneliness and social isolation in 2006 or 2008.

Panel A – Loneliness
|  | 3-Item UCLA Loneliness:<br>50-64 years |  | 3-Item UCLA Loneliness:<br>65-95 years |  |
| --- | --- | --- | --- | --- |
|  | Effect-Size | 95% CI | Effect-Size | 95% CI |
|  | *Mortality (HR) (50-64: N = 5,191; 65-95: N = 5,904)* |  |  |  |
| Mortality | 2.00 | 1.43-2.80 | 1.39 | 1.18-1.64 |
|  | *Prevalent Disability and Disease (IRR) (50-64: N = 4,715; 65-95: N = 4,625)* |  |  |  |
| ADL | 2.20 | 1.85-2.62 | 1.85 | 1.62-2.13 |
| IADL | 2.22 | 1.84-2.67 | 1.81 | 1.58-2.07 |
| Chronic Disease | 1.23 | 1.17-1.29 | 1.12 | 1.08-1.17 |
|  | *Incident Disability and Disease (IRR) (50-64: N = 4,715; 65-95: N = 4,625)* |  |  |  |
| ADL | 1.85 | 1.44-2.37 | 1.53 | 1.27-1.84 |
| IADL | 1.71 | 1.32-2.21 | 1.49 | 1.24-1.78 |
| Chronic Disease | 1.02 | 0.88-1.18 | 1.02 | 0.87-1.19 |

Panel B – Social Isolation
|  | 6-Item Social Isolation:<br>50-64 years |  | 6-Item Social Isolation:<br>65-95 years |  |
| --- | --- | --- | --- | --- |
|  | Effect-Size | 95% CI | Effect-Size | 95% CI |
|  | *Mortality (HR) (50-64: N = 5,258; 65-95: N = 6,044)* |  |  |  |
| Mortality | 1.80 | 1.28-2.54 | 1.32 | 1.15-1.53 |
|  | *Prevalent Disability and Disease (IRR) (50-64: N = 4,773; 65-95: N = 4,715)* |  |  |  |
| ADL | 2.01 | 1.66-2.42 | 1.41 | 1.24-1.61 |
| IADL | 1.89 | 1.55-2.32 | 1.32 | 1.17-1.50 |
| Chronic Disease | 1.17 | 1.11-1.23 | 1.07 | 1.03-1.10 |
|  | *Incident Disability and Disease (IRR) (50-64: N = 4,773; 65-95: N = 4,715)* |  |  |  |
| ADL | 1.84 | 1.41-2.39 | 1.14 | 0.97-1.35 |
| IADL | 1.77 | 1.34-2.35 | 1.16 | 0.99-1.36 |
| Chronic Disease | 1.01 | 0.86-1.19 | 0.96 | 0.84-1.10 |

**Supplemental Table 10.**
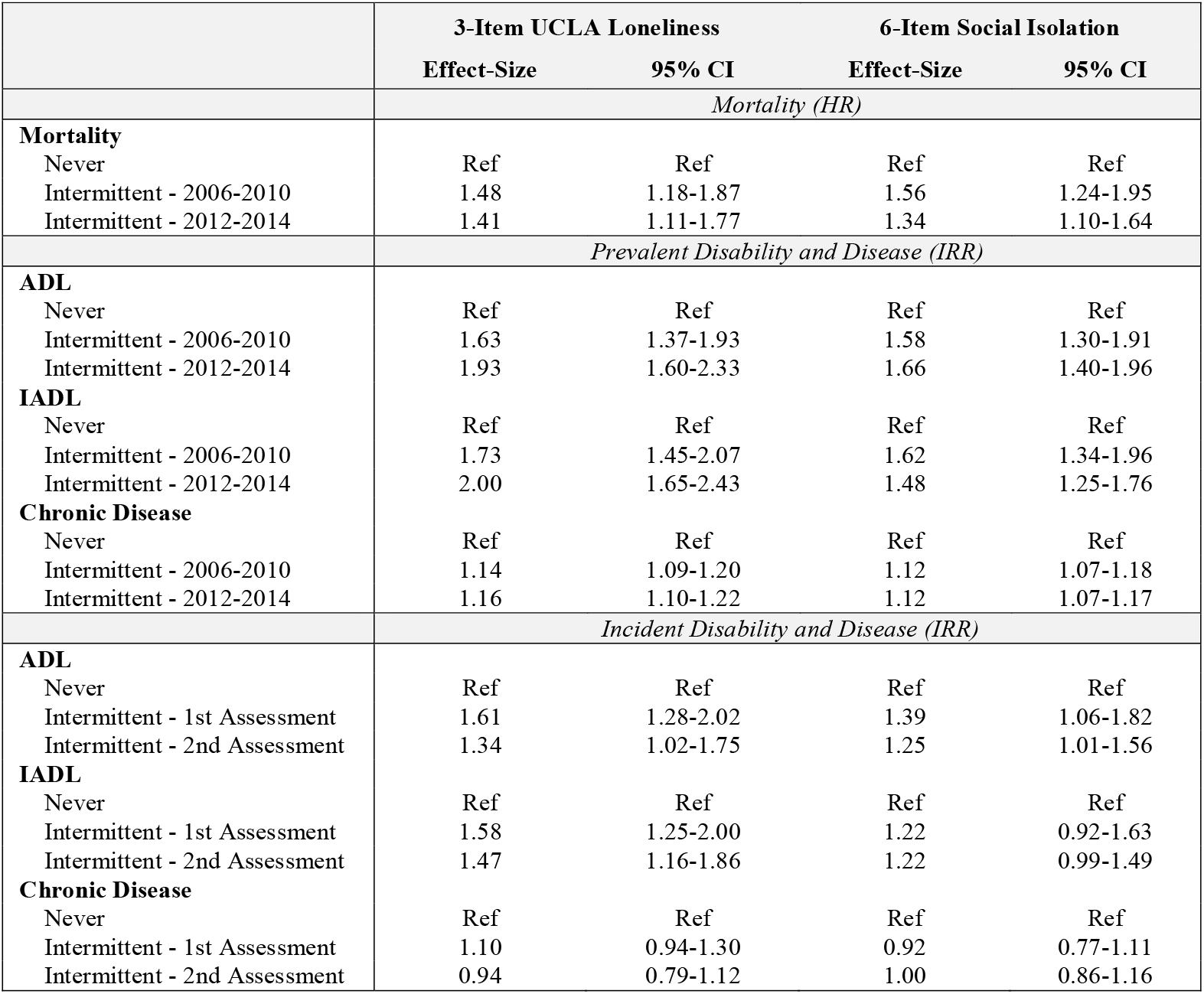
Association of loneliness and social isolation (intermittent vs. never) with deficits in healthy aging, comparing those who were last lonely or socially isolated at their most recent assessment wave and those who were last lonely or socially isolated at previous assessment waves. The table reports effect-sizes for associations of being intermittently lonely or socially isolated compared to never being lonely or socially isolated with deficits in healthy aging. For associations with mortality, effect-sizes are reported as hazard ratios (HR) estimated from Cox proportional hazards models. For prevalent Activities of Daily Living (ADL) disability, Instrumental Activities of Daily Living (IADL) disability, and chronic disease, effect-sizes are reported as incidence rate ratios (IRR) estimated from negative binomial regression models. For all outcomes, effect-sizes are reported separately for those who were last lonely or socially isolated at their most recent assessment wave and those who were last lonely or socially isolated at previous assessment waves. For mortality and prevalent disability and disease, this comparison was between those last lonely or socially isolated in 2012-2014 vs. 2006-2010. For incident disability and disease, this comparison was between those last lonely at their second exposure assessment vs. first exposure assessment. All models included covariate adjustment for age, age-squared, sex, age-sex interactions, race/ethnicity, and a dummy variable coding whether participants were assigned to the subsample of the HRS which first measured loneliness and social isolation in 2006 or 2008.

**Supplemental Figure 1.**
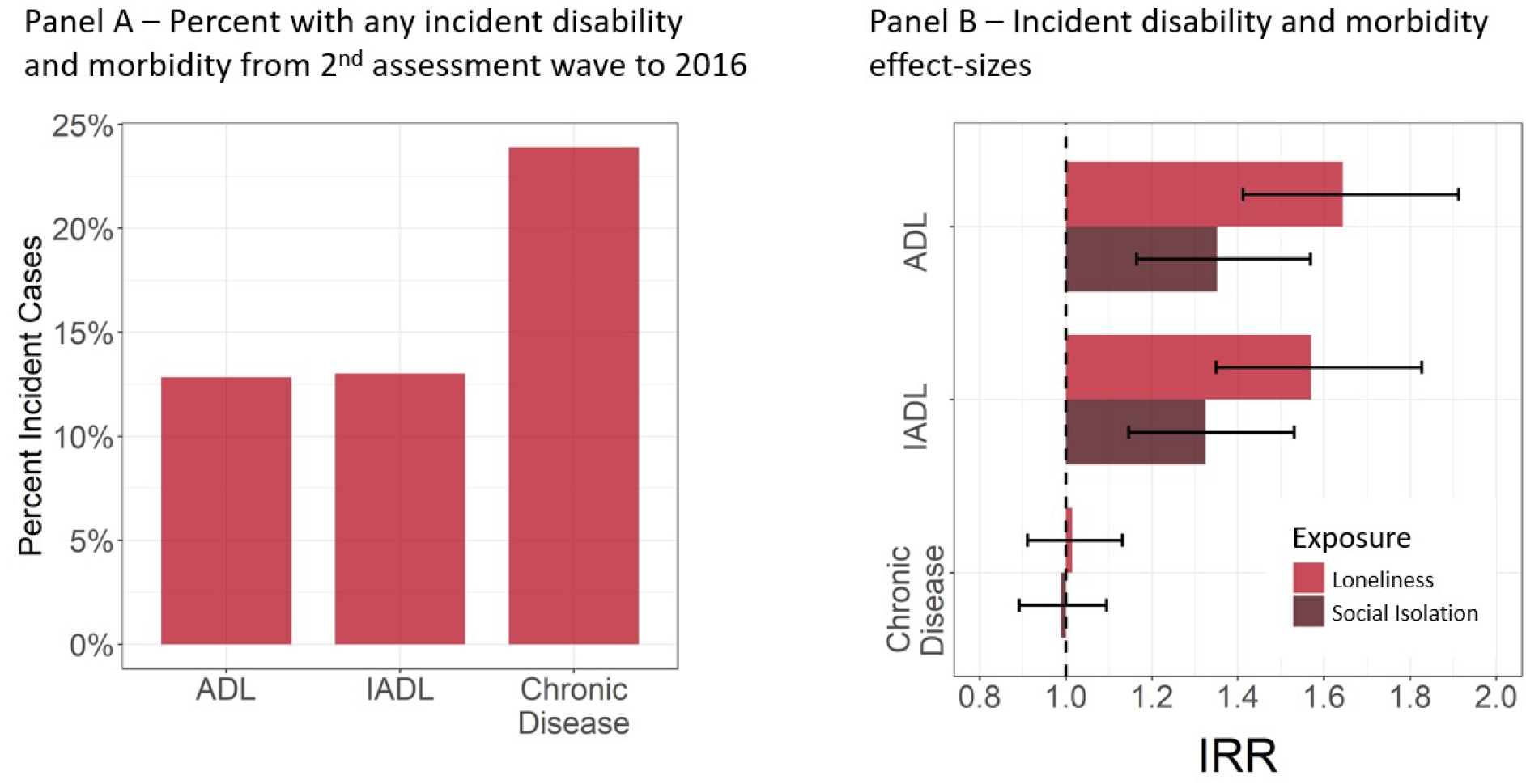
Analysis of incident disability and morbidity. Panel A shows the percent of the analysis sample that reported at least one new ADL limitation, IADL limitation, and chronic disease diagnosis between the second assessment wave and 2016. Panel B shows effect-sizes for analysis of disability and chronic disease (incidence rate ratios (IRR)) from negative binomial regression models including covariate adjustment for age, age-squared, sex, age-sex interactions, race/ethnicity, and a dummy variable coding whether participants were assigned to the subsample of the HRS which first measured loneliness and social isolation in 2006 or 2008. Effect-sizes compare those who ever reported loneliness or social isolation to those who never reported loneliness or social isolation. Red bars represent effect-sizes for loneliness, and brown bars represent effect-sizes for social isolation.

**Supplemental Figure 2.**
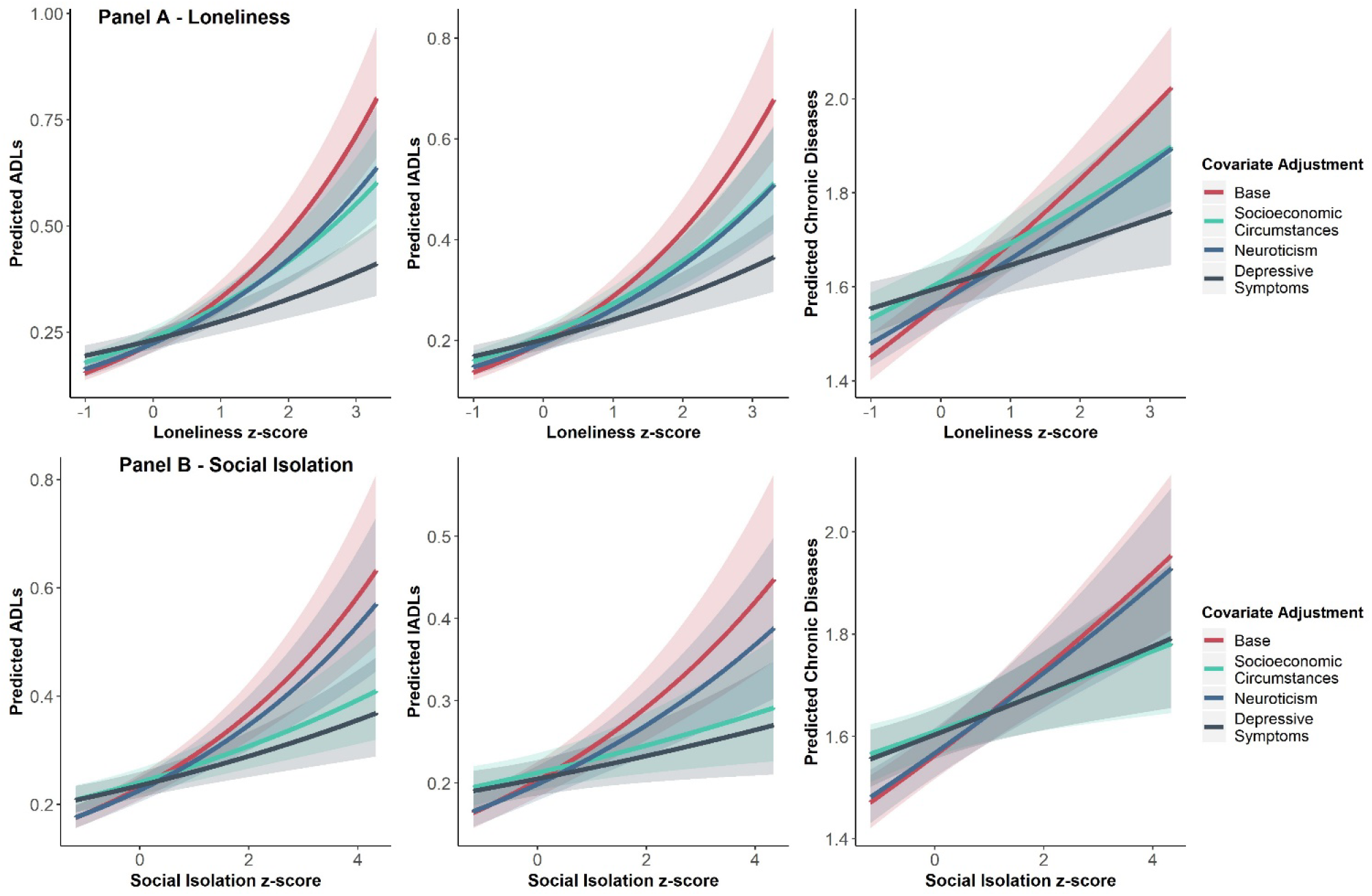
Association of continuous loneliness and social isolation scores with deficits in healthy aging. Panels A and B show the relationship between z-scores (M=0, SD=1) of loneliness and social isolation, respectively, with prevalent disability and disease in 2016. We calculated z-scores based on the average of participants’ loneliness and social isolation scores from 2006-2014. In each plot, z-scores are plotted against the number of predicted Activities of Daily Living limitations (ADLs), Instrumental Activities of Daily Living limitations (IADLs), and chronic disease diagnoses from negative binomial regression models. The base model included covariate adjustment for age, age-squared, sex, age-sex interactions, race/ethnicity, and a dummy variable coding whether participants were assigned to the subsample of the HRS which first measured loneliness and social isolation in 2006 or 2008 (red line). Additional models added covariates for socioeconomic circumstances (green line), neuroticism (blue line), and depressive symptoms (dark blue line). Shaded areas represent 95% confidence intervals.

